# Higher Neighborhood Social Vulnerability is Associated with Lower Life’s Essential 8 Cardiovascular Health Scores: the Coronary Artery Risk Development in Young Adults (CARDIA) Study

**DOI:** 10.64898/2026.05.11.26352953

**Authors:** James Walker, Emily Lam, Daniel Won, Cyanna McGowan, Lucia D. Juarez, Catarina Kiefe, Kiarri N. Kershaw, Hongyan Ning, Donald Lloyd-Jones

**Affiliations:** Department of Preventive Medicine, Northwestern University Feinberg School of Medicine, Chicago, IL; Division of General Internal Medicine and Population Science, Department of Medicine, University of Alabama at Birmingham, Birmingham, AL; Department of Population and Quantitative Health Sciences, University of Massachusetts Chan Medical School, Worcester, MA; Framingham Center for Population and Prevention Science, Boston University Chobanian & Avedisian School of Medicine, Boston, MA

## Abstract

**Background:** Neighborhood social vulnerability may shape cardiovascular health (CVH), but its association with Life’s Essential 8 (LE8), and whether changes in vulnerability track with changes in CVH during midlife, are unclear. We examined cross-sectional and longitudinal associations of the Social Vulnerability Index (SVI) with LE8 and assessed differences by SVI domain, LE8 component, race, and sex.

**Methods:** We analyzed CARDIA participants at Year 15 (Y15; 2000-2001; n = 3,168; mean age 40 years) and Year 30 (Y30; 2015-2016; n = 2,267; mean age 55 years). Residential addresses were geocoded and linked to 2000 and 2016 SVI. Participants were stratified by SVI quartiles. CVH scores were calculated from LE8 metrics (range 0-100; higher is better CVH), excluding sleep. Using multivariable linear regression adjusted for age, sex, race, and educational attainment, we estimated LE8 differences across SVI quartiles and associations of 15-year SVI change/residential mobility with change in LE8. Cox models estimated incident CVD associations.

**Results:** Higher SVI was associated with lower LE8 at both exams. Adjusted Q4 vs Q1 differences in overall LE8 were -5.34 points (95% CI, -6.90 to -3.78) at Y15 and -4.60 points (95% CI, -6.51 to -2.69) at Y30. Among the four SVI domains, SES and household characteristics drove most of the disparity in LE8 scores (Y30 Q4 vs. Q1: SES Δ = –6.98; household Δ = –6.56 points). Component-level differences across quartiles of SVI were largest for nicotine exposure at Y15 (-13.09 points) and physical activity at Y30 (-13.09 points). Changes in SVI and residential mobility were not significantly associated with change in LE8.

**Conclusion:** Higher social vulnerability was associated with significantly lower CVH. Socioeconomic and household factors, along with behavioral gaps in nicotine exposure and physical activity, may be key targets for community-level interventions to improve cardiovascular health equity.

## Introduction

Cardiovascular disease (CVD) remains the leading cause of death in the United States, and large segments of the population still fail to achieve optimal cardiovascular health (CVH).^1^ The American Heart Association’s (AHA) Life’s Essential 8 (LE8) construct assesses an individual’s CVH by scoring their health behaviors and health factors.^1^ Marked deficits in CVH score exist for socioeconomically disadvantaged groups and Black adults.^2^ Higher LE8 scores are strongly associated with lower risks of all-cause and CVD mortality, underscoring the importance of maintaining and improving CVH through primordial and primary prevention.^3,4^

Social determinants of health (SDOH), including income, education, neighborhood context, and structural racism, are strongly associated with CVD risk and outcomes.^5-7^ Individuals with lower socioeconomic position or limited access to health-promoting resources have a higher prevalence of hypertension, diabetes, obesity, and nicotine exposure, worse risk factor control, and greater CVD incidence and mortality.^5-7^ Beyond individual-level SDOH, neighborhood conditions independently shape CVH. Higher neighborhood deprivation has been linked to greater CVD burden and premature mortality, even after accounting for individual-level factors.^8-10^ The Centers for Disease Control and Prevention (CDC)/Agency for Toxic Substances and Disease Registry (ATSDR) Social Vulnerability Index (SVI) is a widely used area-level measure that incorporates a broader set of social and structural risks.^11^ Although developed for disaster preparedness, SVI is increasingly applied in epidemiological research. Higher SVI is associated with greater prevalence of cardiometabolic risk factors, higher odds of self-reported CVD, and worse outcomes across the CVD care continuum.^12,13^ Black and Hispanic populations are more likely to live in high-SVI communities, reflecting the cumulative impact of residential segregation and structural racism.^12,13^ However, most prior SVI work has focused on downstream CVD outcomes or healthcare utilization rather than upstream CVH, and there are limited data directly linking SVI to LE8 scores, particularly in midlife or across race and sex subgroups.

Midlife is a critical period in the life course of CVH, when CVH tends to decline as risk factor trajectories accelerate, yet remain modifiable.^14^ Understanding how neighborhood social vulnerability relates to LE8 in midlife may help identify high-risk communities and inform place-based interventions. We therefore used data from the Coronary Artery Risk Development in Young Adults Study (CARDIA), a long-standing, biracial U.S. cohort followed from young adulthood into midlife, to examine associations between neighborhood SVI and LE8 CVH scores. We evaluated overall SVI and its four domains, characterized differences by race and sex, and assessed patterns of residential “movers” and “stayers” across SVI levels. We hypothesized that higher neighborhood social vulnerability would be associated with lower LE8 scores and secondarily, higher CVD rates at midlife, particularly among Black adults, independent of individual sociodemographic characteristics.

## Methods

### Study Population

CARDIA is a prospective cohort of Black and White adults enrolled in 1985–1986 at ages 18–30 years from 4 U.S. centers (Birmingham, Chicago, Minneapolis, and Oakland). Participants have been re-examined at regular intervals with standardized protocols; additional details are provided in the Supplemental Material. For this analysis, we used data from the Year 15 (Y15; 2000–2001; mean age ∼40 years) and Year 30 (Y30; 2015–2016; mean age ∼55 years) examinations, which correspond to midlife and to available Social Vulnerability Index (SVI) data. The Y15 (n=3,168) and Y30 (n=2,267) analytic samples included participants who attended the exam, had a geocoded residential address, no CVD history prior to Y15, and complete data for LE8 score (excluding sleep, which was not collected in CARDIA Y15), age, sex, race, and educational attainment. Longitudinal analyses were restricted to participants with SVI, LE8, and covariates available at both Y15 and Y30 (n=1,904). Differences between participants who attended only at Y15 versus those included at Y15 and Y30 are included in **Table S4**. All participants provided written informed consent, and institutional review boards at each field center approved the study.

### Neighborhood Social Vulnerability and Residential Change

Neighborhood context was characterized using the CDC/ATSDR SVI, a census tract–level percentile score (0– 1) based on 15 indicators in four domains: socioeconomic status; household composition/disability; minority status/language; and housing/transportation (higher values indicate greater vulnerability). Residential addresses at Y15 and Y30 were geocoded to census tracts and linked to the SVI release closest in time to each exam (2000 and 2016, respectively). Within each exam, overall SVI was categorized into quartiles based on its distribution (quartile 1 [Q1] = least vulnerable; quartile 4 [Q4] = most vulnerable).

To describe residential change, we compared census tracts between Y15 and Y30. Participants who remained in the same tract were classified as “stayers”; those in different tracts were “movers.” Among stayers, we compared tract SVI quartiles at Y15 and Y30 and labeled neighborhoods as improving (shifting to lower quartile), stable, or worsening (shifting to higher quartile). Among movers, we compared origin and destination quartiles and categorized moves as to less vulnerable, same, or more vulnerable neighborhoods.

### Cardiovascular Health (Life’s Essential 8)

Cardiovascular health was measured for each CARDIA participant using the AHA’s LE8 construct (scored 0-100, higher score indicating better CVH), comprising 3 health behaviors (diet, physical activity, nicotine exposure) and 4 health factors (body mass index [BMI], non-high-density lipoprotein cholesterol, blood glucose, blood pressure). The fourth health behavior, sleep, was excluded due to its lack of collection at Y15.

Each metric was scored individually from 0-100 and the overall composite LE8 score was calculated as the mean of the 7 individual metric scores.^1,2^

### Cardiovascular Outcomes

Ascertainment and adjudication of cardiovascular disease (CVD) events in CARDIA have been described previously and are summarized in the Supplemental Material.^15^ For this analysis, we treated incident CVD as a secondary outcome. CVD included the first event of fatal or nonfatal coronary heart disease (definite or probable myocardial infarction or acute coronary syndrome, nonelective coronary revascularization), hospitalized heart failure, peripheral artery disease, transient ischemic event, and stroke.

### Statistical Analysis

We stratified the study sample into 4 groups by neighborhood SVI quartile ranking (0-25%, 25-50%, 50-75% and 75-100%) at Y15 and Y30, respectively. We compared the characteristics across the Y15 SVI quartile groups, using ANOVA for continuous variables or chi-square test for categorical variables. For cross-sectional analyses, multivariable linear regression was used to estimate adjusted mean differences in overall LE8 across overall SVI quartiles at each exam, adjusting for age, sex, race, and educational attainment. Parallel models examined SVI domains and individual LE8 components. For change analyses, we calculated ΔLE8 (Y30 - Y15) and compared mean ΔLE8 across mover/stayer-SVI categories using linear regression adjusted for age, sex, race, and educational attainment.

The rates of incident CVD per 1,000 person-years were calculated using a Poisson regression model. We used multivariable-adjusted Cox proportional hazards models to assess the association between SVI quartile groups and CVD events at Y15 and Y30, taking the lowest quartile (0-25%) as the reference. Model 1 adjusted for age, sex, race, and educational attainment; Model 2 additionally adjusted for baseline LE8. Secondary models examined SVI domains, |ΔSVI|, and mover/stayer-SVI categories. Proportional hazards assumptions were tested using Schoenfeld residuals and log-log survival plots, and no violations were found. A random intercept or the robust sandwich variance estimator was included in the linear or Cox proportional hazards regression model as appropriate to account for the census tract-level clustering effect. All analyses were performed using SAS 9.4 (SAS Institute, Cary, NC).

## Results

### Sample Characteristics

Among 3,168 participants at Y15 and 2,267 at Y30, 1,904 had SVI, LE8, and covariates available at both examinations and were included in longitudinal analyses. At Y15, the analytic sample had mean age 40.2 (standard deviation [SD]: 3.6) years; 45.1% identified as male and 44.8% as Black. Mean overall SVI at Y15 was 0.48 (SD: 0.31) and mean overall LE8 score was 70.9 (SD: 14.8) (**Table 1**). Between Y15 and Y30, 11.9% of participants moved to a different census tract and 88.1% remained in the same tract.

**Table 1.** Baseline Characteristics at Year 15 by Overall Social Vulnerability Index Quartile.

| Characteristic | Overall | Q1 (least vulnerable) | Q2 | Q3 | Q4 (most vulnerable) |
| --- | --- | --- | --- | --- | --- |
| **N** | **ALL**<br>**(n=3168)** | **Q1 (n=793)** | **Q2 (n=791)** | **Q3 (n=792)** | **Q4 (n=792)** |
| **Age, years (SD)** | 40.2 (3.6) | 40.8 (3.3) | 40.2 (3.5) | 40.1 (3.7) | 39.8 (3.8) |
| **Male sex, n (%)** | 1430<br>(45.1%) | 379 (47.8%) | 387 (48.9%) | 331 (41.8%) | 333 (42.0%) |
| **Black, n (%)** | 1419<br>(44.8%) | 133 (16.8%) | 238 (30.1%) | 387 (48.9%) | 661 (83.5%) |
| **Maximum years of education (SD)** | 15.8 (2.6) | 16.8 (2.4) | 16.1 (2.6) | 15.6 (2.6) | 14.8 (2.5) |
| **High school or less, n (%)** | 428<br>(13.5%) | 37 (4.7%) | 91 (11.5%) | 117 (14.8%) | 183 (23.1%) |
| **Some college/associate, n (%)** | 923<br>(29.1%) | 151 (19.0%) | 201 (25.4%) | 256 (32.3%) | 315 (39.8%) |
| **College graduate or higher, n (%)** | 1817<br>(57.4%) | 605 (76.3%) | 499 (63.1%) | 419 (52.9%) | 294 (37.1%) |
| **Annual family income ≥\$45,000, n (%)** | 1885<br>(60.1%) | 665 (84.4%) | 536 (68.2%) | 431 (55.1%) | 253 (32.4%) |
| **Financial strain, n (%)** |  |  |  |  |  |
| **Difficulty paying for basics** | 580<br>(18.3%) | 78 (9.8%) | 106 (13.4%) | 169 (21.4%) | 227 (28.8%) |
| **Difficulty paying for medical care** | 599<br>(19.0%) | 97 (12.3%) | 116 (14.7%) | 159 (20.2%) | 227 (28.8%) |
| **Overall SVI, 0–1 (SD)** | 0.48 (0.31) | 0.10 (0.06) | 0.33 (0.07) | 0.62 (0.09) | 0.88 (0.06) |
| **SVI domains, 0–1 (SD)** |  |  |  |  |  |
| **Socioeconomic status** | 0.43 (0.30) | 0.12 (0.10) | 0.27 (0.15) | 0.52 (0.17) | 0.83 (0.13) |
| **Household composition/disability** | 0.45 (0.32) | 0.15 (0.12) | 0.29 (0.20) | 0.51 (0.26) | 0.85 (0.14) |
| **Minority status/language** | 0.57 (0.29) | 0.35 (0.24) | 0.50 (0.26) | 0.68 (0.23) | 0.75 (0.22) |
| **Housing/transportation** | 0.50 (0.29) | 0.18 (0.16) | 0.46 (0.22) | 0.64 (0.22) | 0.73 (0.19) |
| **Overall LE8 score, 0–100 (SD)** | 70.9 (14.8) | 75.5 (13.6) | 72.9 (14.2) | 70.6 (14.5) | 64.4 (14.5) |
| **Unadjusted Y15 LE8 behaviors, 0–100 (SD)** |  |  |  |  |  |
| **Diet** | 42.9 (32.9) | 50.3 (31.9) | 46.9 (32.5) | 43.8 (33.5) | 30.8 (30.4) |
| **Physical activity** | 73.5 (38.6) | 77.2 (36.0) | 75.0 (37.9) | 72.4 (39.0) | 69.3 (41.0) |
| **Nicotine exposure** | 70.1 (39.5) | 78.0 (34.3) | 74.3 (36.8) | 70.6 (39.0) | 57.7 (44.3) |
| **Y15 LE8 factors, 0–100 (SD)** |  |  |  |  |  |
| **Body mass index** | 62.7 (33.9) | 70.1 (30.5) | 66.4 (32.2) | 60.6 (34.4) | 53.6 (35.8) |
| **Non-HDL cholesterol** | 72.7 (29.4) | 72.1 (29.3) | 71.8 (29.4) | 73.8 (29.1) | 73.1 (29.7) |
| **Blood glucose** | 95.3 (14.8) | 96.8 (12.0) | 95.5 (14.4) | 95.6 (14.4) | 93.1 (17.7) |
| **Blood pressure** | 78.8 (30.5) | 84.3 (25.7) | 80.7 (29.7) | 77.5 (31.2) | 72.9 (33.6) |

**Note:** Values are mean (SD) for continuous variables and n (%) for categorical variables. Participants were grouped by quartile of overall Social Vulnerability Index (SVI), with Q1 representing the least vulnerable and Q4 the most vulnerable neighborhoods. Sleep score was not included in the Life's Essential 8 (LE8) score because sleep data were unavailable at the Year 15 exam. Accordingly, overall LE8 was calculated as the mean of the remaining 7 component scores (0–100). For financial strain variables, “yes” was defined as anyone who answered “very hard,” “hard,” or “somewhat hard,” while “no” was defined as anyone who answered “not very hard.” Denominators vary because of missing data for income/financial strain variables.

### Neighborhood Vulnerability by Race and Sex

Neighborhood social vulnerability differed substantially by race but minimally by sex. At Y15, mean overall SVI was 0.64–0.66 among Black participants and 0.33–0.35 among White participants, with similar gaps at Y30 (**Table S2**). In the SES domain, mean SVI was 0.59–0.61 vs 0.28–0.30 for Black vs White participants at Y15, with a similar gap at Y30, indicating that Black adults in CARDIA, on average, resided in neighborhoods with substantially higher socioeconomic vulnerability than White adults. Within each race, differences in overall and domain-specific SVI by sex were small.

### Association of Neighborhood Social Vulnerability With LE8 Score

Higher neighborhood social vulnerability was consistently associated with lower LE8 scores at both midlife examinations. After adjustment for age, sex, race, and educational attainment, mean LE8 was lower with higher neighborhood vulnerability overall, with the clearest gradient at Y15 (**Figure 1**). At Y15, participants in the most vulnerable quartile (Q4) had 5.34-point lower LE8 (95% CI: –6.90 to –3.78) scores than those in the least vulnerable quartile (Q1). At Y30, Q4–Q1 difference was –4.60 points (95% CI: –6.51, –2.69), with lower LE8 scores overall at higher levels of neighborhood vulnerability (**Figure 1**). In a sensitivity analysis excluding race from covariate adjustment, the inverse association was stronger, particularly for Q4, with Q4 vs Q1 differences of –8.54 points (95% CI: –9.99 to –7.10) at Y15 and –8.48 points (95% CI: –10.27 to –6.69) at Y30 (**Figure S2**).

**Figure 1:**
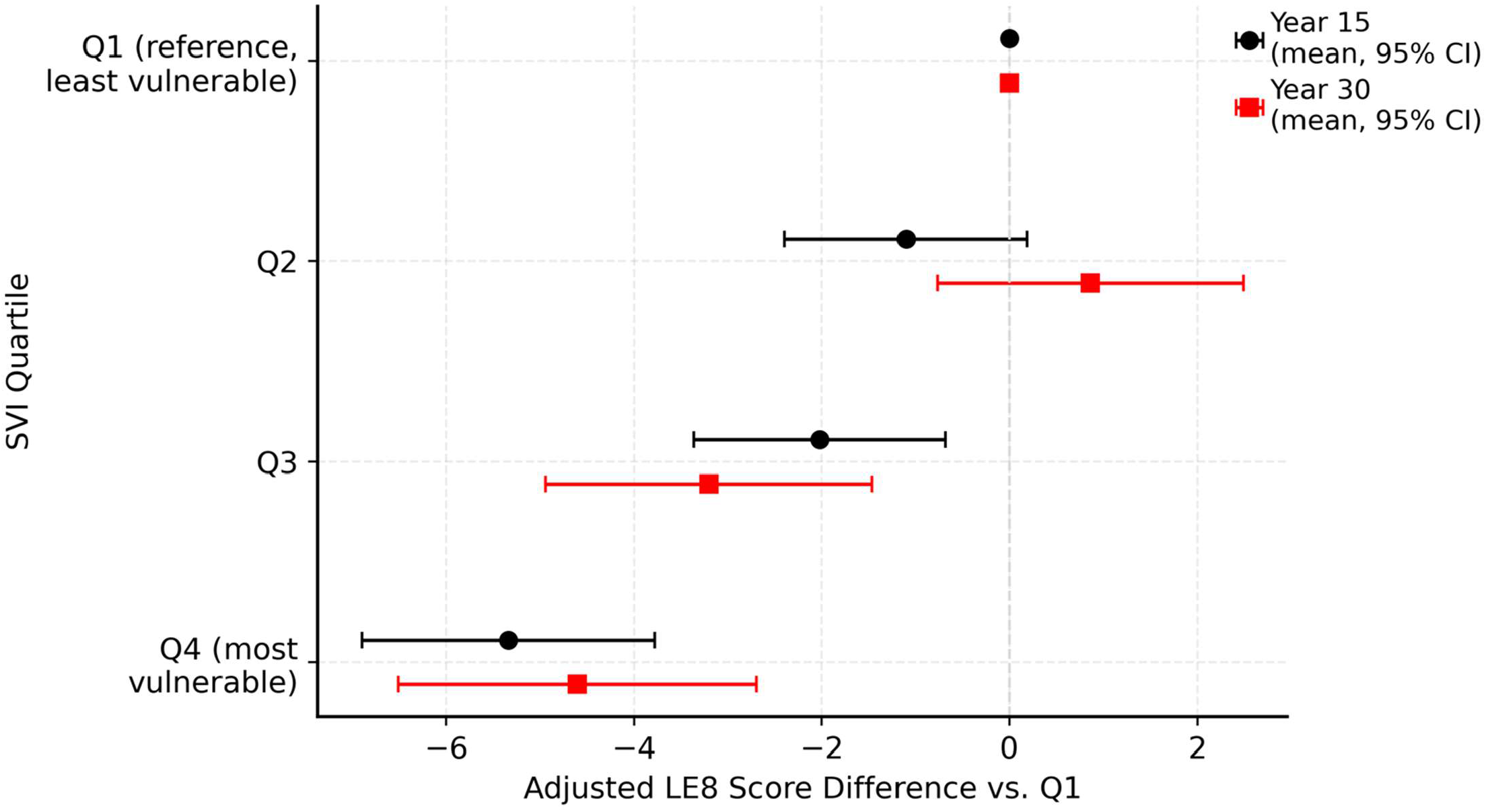
Adjusted cross-sectional differences in overall LE8 score by overall SVI quartile, Q2–Q4 vs. Q1 (reference, least social vulnerability) in CARDIA Y15 and Y30. Adjusted mean differences vs. Q1 are shown (adjusted for age, sex, race, and educational attainment), and error bars show 95% CIs. More negative values indicate lower cardiovascular health.

When SVI domains were examined separately, socioeconomic status and household composition were most strongly associated with CVH (**Table S1**). At Y15, adjusted LE8 scores were 4.72 and 5.02 points lower in the most vs least vulnerable quartiles of the SES and household domains, respectively, compared with 2.65 and 1.98 points lower for the minority status/language and housing/transportation domains. At Y30, gaps for the SES and household domains widened to 6.98 and 6.56 points, whereas differences were less than two points for the minority status/language and housing/transportation domains.

### LE8 Component Scores Across SVI Quartiles

Disparities in overall LE8 scores by SVI quartiles were driven largely by behavioral components (**Figure 2**). Across the extremes of overall SVI, the largest gaps were observed for nicotine exposure and physical activity scores. At Y15, the Q4–Q1 difference was –13.09 points for nicotine exposure and –7.38 points for physical activity; at Y30, the corresponding differences were –8.43 and –13.09 points, respectively. For other metrics, Q4–Q1 differences were generally smaller and less consistent, with body mass index showing moderate deficits and non-HDL cholesterol showing little consistent difference across SVI quartiles. Associations of LE8 with SVI change quartiles were also not statistically significant (**Table S3**).

**Figure 2:**
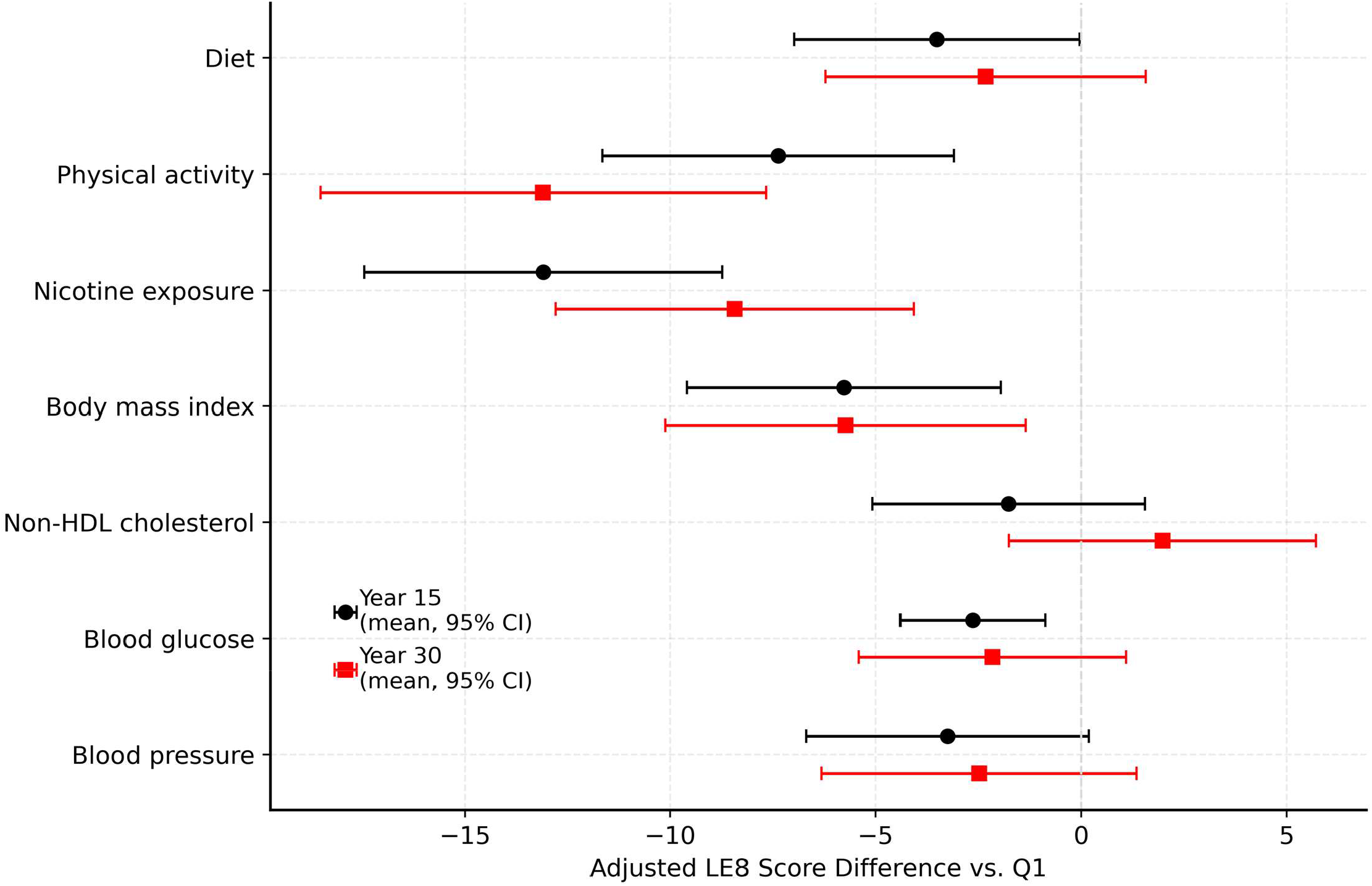
Adjusted cross-sectional difference in LE8 component scores, Q4 vs. Q1 of overall SVI, CARDIA Y15 and Y30. Negative scores indicate worse cardiovascular health.

### Neighborhood Vulnerability, LE8 Trajectories, and Incident CVD

Across mover/stayer and SVI-change categories, overall CVH worsened between Y15 and Y30, with similar declines observed in all groups **(Figure 3)**. Mean LE8 declined by approximately 5–7 points over 15 years within each mover/stayer–SVI group, and differences in ΔLE8 across these groups and across quartiles of continuous ΔSVI were small and not statistically significant, suggesting that midlife changes in neighborhood vulnerability were not strongly related to within-person changes in LE8 during this period.

**Figure 3:**
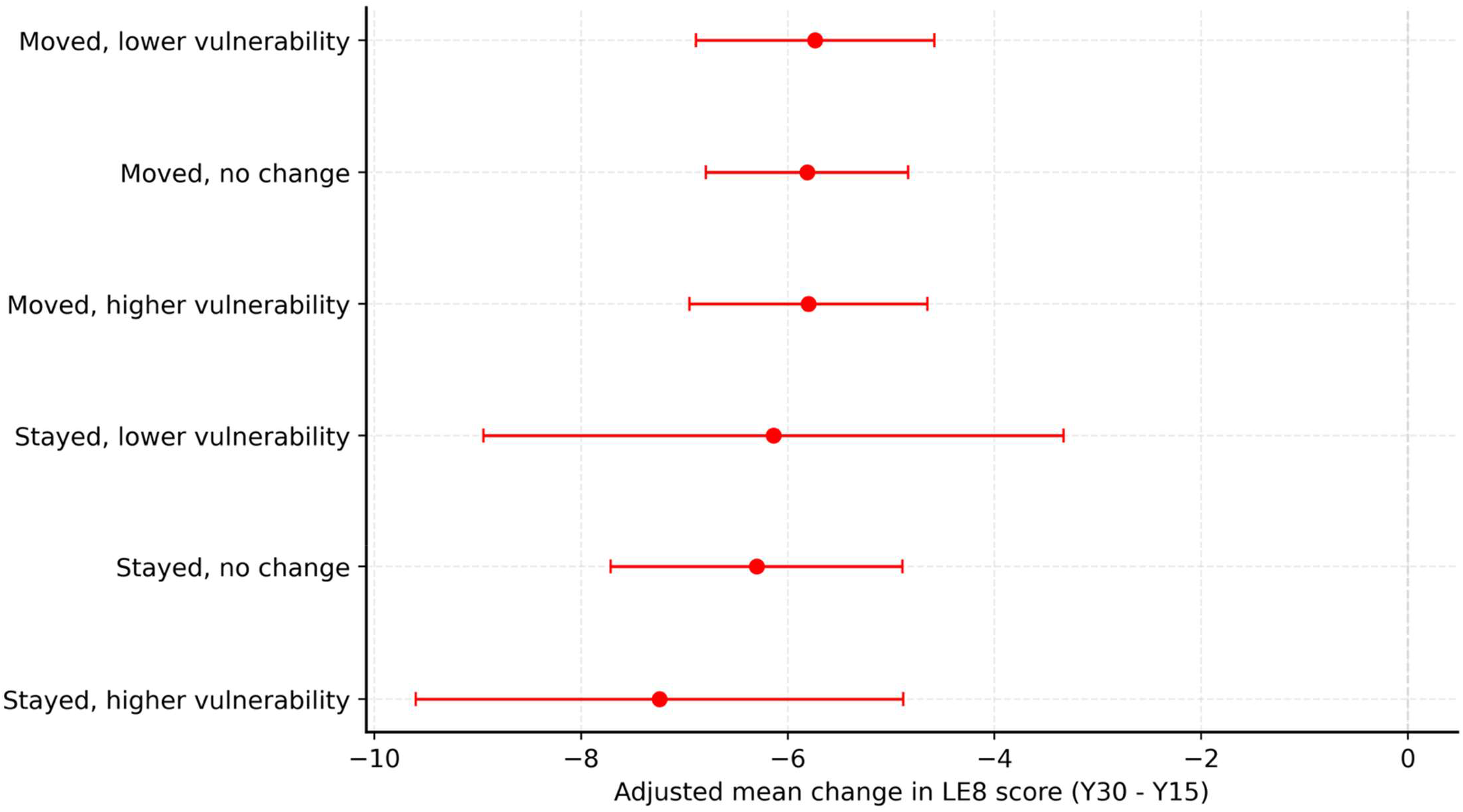
Change in LE8 from Y15 to Y30 by residential mobility and change in neighborhood SVI quartile. Points indicate adjusted mean change in LE8. Negative scores indicate worse cardiovascular health, and horizontal bars show 95% confidence intervals. At Y30, 88.1% of participants were still living in their Y15 census tract, whereas 11.9% had moved to a different census tract.

By contrast, cross-sectional neighborhood vulnerability at Y15 was associated with incident CVD (**Table 2**). At Y15, there were 49, 49, 46, and 106 incident CVD events in Q1 through Q4 of SVI, respectively. Unadjusted CVD event rates per 1,000 person-years at Y15 were 2.84 (95% CI: 2.15, 3.76), 2.85 (2.15, 3.77), and 2.70 (2.02, 3.61) in SVI quartiles Q1, Q2, and Q3 respectively, compared to 6.41 (5.29, 7.77) in Q4; the Q4–Q1 difference was statistically significant (p<0.01), whereas differences for Q2 and Q3 vs Q1 were not. In Cox models adjusted for age, sex, race, and educational attainment, participants in Q4 had nearly twice the hazard of CVD compared with Q1 (HR 1.92; 95% CI: 1.30, 2.82); further adjustment for baseline LE8 attenuated but did not eliminate this association (HR 1.52; 95% CI: 1.04, 2.23) **(Table 2)**. At Y30, the corresponding event counts were 13, 8, 22, and 21 in Q1 through Q4, respectively. CVD event rates were more heterogeneous across SVI quartiles (3.03, 1.86, 5.17, and 4.93 per 1,000 person-years for Q1 to Q4, respectively), and none of the rate differences vs Q1 was statistically significant. Adjusted hazard ratios for Q2–Q4 vs Q1 at Y30 were imprecise with wide confidence intervals crossing 1, and there was no clear trend.

**Table 2.** Incident CVD Rates and Adjusted Hazard Ratios by Social Vulnerability Index Quartile at CARDIA Y15 and Y30.

| CARDIA year | SVI quartile | # of CVD events | CVD rate per 1000 PY (95% CI) | P value for rate vs Q1 | HR (95% CI), Model 1* | P value (Model 1) | HR (95% CI), Model 2** | P value (Model 2) |
| --- | --- | --- | --- | --- | --- | --- | --- | --- |
| Y15 | Q1 | 49 | 2.84 (2.15–3.76) | ref | 1.00 (ref) | ref | 1.00 (ref) | ref |
| Y15 | Q2 | 49 | 2.85 (2.15–3.77) | 0.99 | 0.95 (0.63–1.41) | 0.80 | 0.87 (0.58–1.32) | 0.53 |
| Y15 | Q3 | 46 | 2.70 (2.02–3.61) | 0.80 | 0.88 (0.59–1.32) | 0.55 | 0.79 (0.53–1.19) | 0.26 |
| Y15 | Q4 | 106 | **6.41 (5.29–7.77)** | **<0.01** | **1.92 (1.30–2.82)** | **<0.01** | **1.52 (1.04–2.23)** | **0.03** |
| Y30 | Q1 | 13 | 3.03 (1.76–5.21) | ref | 1.00 (ref) | ref | 1.00 (ref) | ref |
| Y30 | Q2 | 8 | 1.86 (0.93–3.72) | 0.28 | 0.61 (0.26–1.45) | 0.27 | 0.63 (0.26–1.50) | 0.30 |
| Y30 | Q3 | 22 | 5.17 (3.41–7.86) | 0.13 | 1.58 (0.76–3.29) | 0.22 | 1.47 (0.70–3.09) | 0.30 |
| Y30 | Q4 | 21 | 4.93 (3.21–7.56) | 0.17 | 1.39 (0.62–3.13) | 0.42 | 1.24 (0.54–2.86) | 0.61 |

**Note:** CVD rates are expressed as events per 1,000 person-years (PY) with 95% confidence intervals (CIs); p values for rates compare each quartile with Q1. Hazard ratios (HRs) are from Cox proportional hazards models comparing quartiles Q2–Q4 with Q1 (reference). \*Model 1 adjusted for age, sex, race, and educational attainment. \*\*Model 2 additionally adjusted for baseline Life's Essential 8 (LE8) score at the corresponding examination. Ref indicates reference group. Bold indicates significant p values.

## Discussion

In this prospective analysis of a biracial cohort followed from young adulthood into midlife, we observed a consistent inverse association between neighborhood social vulnerability and cardiovascular health. Participants residing in the most vulnerable neighborhoods had LE8 scores approximately 5 points lower than those in the least vulnerable neighborhoods. Recent data suggest that lower LE8 score is significantly associated with higher mortality and incident CVD.^4,16^ The disparity in CVH across SVI was most evident in health behaviors (nicotine exposure and physical activity) rather than clinical factors (BMI, non-HDL cholesterol, blood glucose, or blood pressure). This is consistent with the concept of “obesogenic environments” and “tobacco swamps,” where high-vulnerability communities often have structural barriers that clinical care alone cannot easily surmount (e.g., lack of safe green spaces for recreation or high densities of tobacco retailers).^17,18^

Our findings extend prior research by analyzing which aspects of vulnerability drive the gap in CVH. While previous studies have demonstrated associations between the SVI and downstream outcomes such as premature cardiovascular mortality and heart failure readmissions, fewer have examined the upstream association with CVH in younger individuals.^19,20^ We found that SES and household composition domains were the primary drivers of LE8 disparities, whereas the minority status and housing domains showed weaker independent associations. This suggests that the specific material deprivation of a community (e.g., poverty, unemployment, and single-parent household density) may be more linked to CVH behaviors than the built environment features captured in the housing domain.

However, this does not negate the role of structural racism. We observed that Black participants were overwhelmingly concentrated in the higher quartiles of overall SVI and the SES domain. This extreme divergence in residential context, likely a legacy of redlining and discriminatory housing policies, means that the “average” neighborhood experience for Black adults in midlife is fundamentally different from that of White adults, creating a structural headwind against maintaining or improving cardiovascular health.^21^ Consistent with this interpretation, removing race from the adjustment set strengthened the inverse association between SVI and LE8, suggesting that racial differences in neighborhood vulnerability account for an important share of the observed population-level disparity in cardiovascular health.

We hypothesized that moving to a lower-vulnerability neighborhood would be associated with improvements in LE8, yet we found no significant change in LE8 scores among movers compared to stayers. Several factors may explain this. First, the “lag time” for health behaviors to translate into measurable score changes may exceed the observation window; moving is a disruptive life event that may temporarily hinder health maintenance before long-term benefits accrue. Second, “sorting” may occur, where individuals move to neighborhoods that are only marginally different in vulnerability or where they carry their established health behaviors with them.^22^ Finally, CVD risk is cumulative. As shown in our prior work regarding cumulative LE8 exposure, the physiological damage of residing in a high-vulnerability environment during young adulthood may not be immediately reversible by a midlife change in address.^14^ This suggests that place-based interventions may need to be sustained over decades, rather than relying on residential mobility as a solution.

Our analysis of incident CVD revealed a strong association between high social vulnerability and premature CVD events at Year 15, but a more heterogeneous pattern at Year 30. This apparent attenuation may reflect a survival bias (or “healthy survivor” effect). However, the number of incident CVD events after the Y30 examination was modest across SVI quartiles given the shorter follow-up than after Y15, resulting in wide confidence intervals and limited power to detect associations at Y30. Therefore, the attenuated findings at Y30 should be interpreted cautiously. Participants living in the most vulnerable neighborhoods experienced significantly higher rates of premature events prior to the Year 30 exam, effectively removing the most susceptible individuals from the “at-risk” pool for the later cross-sectional analysis. This interpretation is supported by the stronger associations observed for Y15 neighborhood vulnerability in the adjusted hazard models and highlights the importance of intervening early in the life course, before the most vulnerable populations succumb to premature morbidity.

## Limitations

Limitations include the observational nature of the study, which precludes causal inference. There is a possible attrition bias due to the loss to follow-up of some CARDIA participants. While we adjusted for individual-level age, sex, race, and educational attainment, residual confounding remains possible. Additionally, the SVI is a composite measure; while we analyzed domains separately, specific neighborhood features (e.g., food swamps vs. food deserts) are not distinguished. CARDIA sleep data were not collected until Year 20 and thus sleep scores were not factored into the analysis. LE8 scores and the mover analyses relied on data collected at two time points, which does not capture the full variation in CVH and neighborhood mobility experienced by participants over the 15-year period. Further, the number of incident CVD events after the Y30 examination was relatively small, which limited precision and power for the Y30 event-rate and hazard ratio analyses. Finally, the cohort’s inclusion of only two races, Black and White, limits its generalizability.

## Conclusion

Neighborhood social vulnerability, particularly in socioeconomic and household domains, is significantly associated with lower cardiovascular health in midlife, largely manifesting in deficits in physical activity and nicotine avoidance. These structural disadvantages disproportionately affect Black adults and contribute to disparities in premature cardiovascular disease. The lack of change in LE8 among movers suggests that improving cardiovascular equity requires more than individual residential mobility; it may require sustained, place-based investment to transform high-vulnerability communities into health-promoting environments.

## Data Availability

Deidentified CARDIA data used in this analysis are available to qualified investigators through the National Heart, Lung, and Blood Institute Biologic Specimen and Data Repository Information Coordinating Center (BioLINCC), subject to its data request and approval procedures. Some restricted variables, including geocoded residential information used for neighborhood linkage, may require additional approval through the CARDIA Coordinating Center. The CDC/ATSDR Social Vulnerability Index data used for neighborhood exposure assignment are publicly available.

https://www.atsdr.cdc.gov/place-health/php/svi/svi-data-documentation-download.html

https://biolincc.nhlbi.nih.gov/studies/cardia/

## Acknowledgements

The Coronary Artery Risk Development in Young Adults Study (CARDIA) is conducted and supported by the National Heart, Lung, and Blood Institute (NHLBI) in collaboration with the University of Alabama at Birmingham (75N92023D00002 & 75N92023D00005), Northwestern University (75N92023D00004), University of Minnesota (75N92023D00006), and Kaiser Foundation Research Institute (75N92023D00003). This manuscript has been reviewed by CARDIA for scientific content.

